# Is the self-reporting of mental health problems sensitive to public stigma towards mental illness? A comparison of time trends across English regions (2009-19)

**DOI:** 10.1101/2022.08.08.22278542

**Authors:** Thierry Gagné, Claire Henderson, Anne McMunn

## Abstract

**Purpose:** The prevalence of mental health problems has rapidly increased over time. The extent to which this captures changes in self-reporting due to decreasing stigma is unclear. We explore this by comparing time trends in mental health and stigma-related indicators across English regions.

**Methods:** We produced annual estimates of self-reported mental disorders (SRMDs) across waves of the Health Survey for England (2009-18, n = 78,226) and three stigma-related indicators (knowledge, attitudes, and intended behaviour) across waves of the Attitudes Towards Mental Illness survey (2009-19, n = 17,287). Differences in trends were tested across nine Government Office Regions using linear models, adjusting for age, sex, ethnicity, marital status, and social class.

**Results:** In 2009, SRMDs did not vary by region (*p* = .916) whereas stigma-related indicators did (*p* < .001), with London having the highest level of stigma and the North East having lowest level of stigma. Between 2009 and 2018-19, SRMDs increased and stigma-related indicators improved at different rates across regions (SRMDs *p* = .024; stigma-related indicators *p* < .001). London reported the lowest increase in SRMDs (+0.3 percentage point per year) yet among the largest improvements in attitudes and intended behaviour across regions.

**Conclusions:** Improvements in attitudes towards mental illness did not mirror changes in self-reported mental health problems across English regions over the past decade. The findings do not support the argument that changes in public stigma, at least when defined at this regional scale, have been driving the increase in self-reported mental health indicators in recent years.

## 1. INTRODUCTION

Multiple studies in the United Kingdom (UK) have found a large increase in the prevalence of mental health problems over the past two decades that intensified in the years leading to the COVID-19 pandemic, particularly among young people [1–4]. A key inferential issue has been the extent to which these time trends capture meaningful increases in the burden of mental health or changes in self-report practices. In particular, many point to increased awareness and reduced stigma over time as an alternative hypothesis for changes in individuals’ propensity to self-report mental health problems [4]. The experience of stigma has already been linked to feelings of distress and shame, and a lower capacity to seek treatment [5]. Discussions about its impact on time trends, however, have often been restricted to limitations sections, and no study that we know of has sought to disentangle the extent to which this may be true.

Stigma around mental illness operates at individual, interpersonal, and societal levels [6]. An appropriate design to test this hypothesis may look at changes in self-reported problems and attitudes towards mental health at the micro- (e.g., household), meso- (e.g., network, community), and macro-level (e.g. region, country) scale over time. While attitudes towards mental health have been measured across a number of UK surveys, it is relatively difficult to derive a comprehensive portrait of trends because attitudes have been measured either once or with different items over time [7–9]. While surveys on attitudes have been run in other countries and trends have been derived using meta-analytic methods, this work remains limited by the lack of repeated measurements, non-representative sampling methods, and/or small sample sizes [10–12].

One key exception is the national Attitudes Towards Mental Illness (AMI) survey, an annual survey of approx. 1,700 people repeated multiple times since 1994 across England [7, 8, 13]. Supporting the argument that attitudes towards mental health have improved over time, studies using AMI data found that both knowledge and attitudes towards people with mental illness improved over the last two decades, together with a reduced desire for social distance from people with mental illness [13, 14]. Further analyses in the AMI survey found that changes in these outcomes over time varied by age, improving more rapidly in younger adults, and by region, improving more rapidly in the region of London [13, 14]. While regional differences have declined over time, they remained significant in 2017-19, with London continuing to show a higher level of stigma around mental illness [13]. Analyses in the 2014 HSE dataset, which included a one-time module on attitudes towards mental illness, also supported the idea that regional differences remained meaningful even after taking into account differences in inhabitants’ age, sex, ethnicity, education, and income across regions [15].

To examine the “self-report bias” hypothesis, we therefore propose to examine how self-reported mental health problems may vary as a function of changes in public stigma towards mental illness across English regions over time. We examine this in four steps: 1) reporting estimates of self-reported mental disorders and stigma-related indicators at two points over the last decade (2009 and 2017-19) and testing cross-sectional differences across regions at both time points; 2) estimating annual change rates in each region and testing differences in these rates between regions; 3) testing the role of socio-demographic composition (and its change over time) through statistical adjustment; 4) testing the relevance of region as an analytic scale by estimating the proportion of the variance in these variables that may be explained at the region level (i.e., context) [16]. If the “self-report bias” hypothesis holds, we expect to see larger increases in the prevalence of self-reported mental health problems in regions that also had larger improvements in stigma-related indicators over time.

## 2. METHODS

### 2.1 Data

We used two datasets to compare time trends over the past decade: the Health Survey for England (HSE) (2009-18) and Attitudes Towards Mental Illness (AMI) survey (2009-19).

The HSE represents a series of annual surveys started in 1991 and designed to monitor trends in the nation’s health [17]. HSE adult samples over the past decade averaged 8,000 participants aged 16+, except in 2009 (*n* = 4,645) where it was made smaller to include a larger sample of children. As a reference point, the estimated individual response rates in 2009 and 2018 were 61% and 54%. Sampling errors are calculated by integrating cluster and stratification variables into the analyses.

The AMI survey has been carried out in England every year from 2008 to 2017, and once again in 2019, by the agency Kantar TNS [13]. There are approximately 1,700 participants aged 16+ for each survey year. A quota sampling frame was used to ensure that the survey sample included sufficient numbers of participants across English regions, with sample points selected by a random location methodology. Sampling errors are calculated on the assumption of a simple random sampling method.

### 2.2 Measures

#### Mental health outcome

We used one measure to capture self-reported mental disorders (SRMDs) in the HSE, derived by the data management team based on open responses with regard to longstanding health conditions (Yes / No). The filter question was *“Do you have any long-standing illness, disability or infirmity? By long-standing I mean anything that has troubled you over a period of time, or that is likely to affect you over a period of time?”* in 2009-11 and *“Do you have any physical or mental health conditions or illnesses lasting or expected to last 12 months or more?”* in 2012-18. The coding frame used in the derived variable available for the time period included both mental illnesses (e.g., alcoholism, drug addiction, anxiety, depression, schizophrenia) and handicaps (see conditions in Supplementary Table 1). While HSE was run in 2019, the type of longstanding health conditions was not asked that year.

#### Stigma-related indicators

Based on the assumption that stigma can be conceptualized as comprised of three constructs, i.e., knowledge (ignorance), attitudes (prejudice), and behaviour (discrimination), we used three separate measures to capture changes in public stigma over time in the AMI survey (see items in Supplementary Table 2) [13]:

1. The Mental Health Knowledge Schedule (MAKS) scale was developed in the late 2000’s to assess stigma-related knowledge about mental health problems among the general public [18]. The scale is composed of six Likert-type items asking whether participants agree with knowledge-related statements (e.g., “If a friend had a mental health problem, I know what advice to give them to get professional help”). MAKS items’ response scale vary from 1 – Strongly Disagree to 5 – Strongly Agree and include a “I do not know” option. Item responses were recoded into a composite score ranging from 6 to 30, with the “I do not know” responses recoded as the mid-point “Neither agree nor disagree” (Cronbach’s alpha in the 2019 sample = .60).
2. The 27-item Community Attitudes towards Mental Illness (CAMI) scale is shortened version of a longstanding item battery initially developed to capture neighbourhood opposition to community-based mental health facilities in the 1970s [19]. CAMI items’ response scale vary from 1 – Strongly Disagree to 5 – Strongly Agree, and item responses were recoded into a composite score ranging from 27 to 135 (Cronbach’s alpha in the 2019 sample = .88).
3. The Reported and Intended Behaviour Scale (RIBS) for desire for social distance was developed in the late 2000’s based on the Social Distance Scale to assess intended stigmatising and discriminatory behaviours towards people with mental health problems [20]. The scale is composed of four Likert-type items asking whether participants agree that they would be willing to live with, work with, live nearby, and be friends with someone with a mental health problem. RIBS items’ response scale vary from 1 – Strongly Disagree to 5 – Strongly Agree, and item responses were recoded into a composite score ranging from 4 to 20 (Cronbach’s alpha in the 2019 sample = .86).

#### Region and covariates

Region was defined using the nine-category Government Office Region classification (i.e. North East, North West, Yorkshire and the Humber, East Midlands, West Midlands, East of England, London, South East, and South West). The robustness of regional differences was tested using five covariates: 1) age (16-24, 25-44, 45-64, and 65+); 2) sex (male or female); 3) ethnicity (White, Black, Asian, and Other); 4) marital status (never married, married, and separated, divorced or widowed); 5) social class of the person responsible for the household. Social class in the AMI was defined using the Market Research Society’s classification system (AB – Managerial & professional occupations, C1 – Skilled non-manual occupations, C2 – Skilled manual occupations, DE – Unskilled manual occupations and not in paid work). Social class in the HSE was defined using the National Statistics Socioeconomic classification (1 – Managerial & professional occupations; 2 – Intermediate occupations; 3 – Routine and manual occupations, 4 – Not in paid work) [21].

### 2.3 Statistical analysis

We first report the distribution of SRMDs and stigma-related indicators across annual waves at the national level and across the nine regions. We tested cross-sectional regional differences in variables in the 2009 and 2018 samples for the HSE, and in the 2009 and 2017-2019 samples for the AMI survey. We then report unadjusted and adjusted estimates of annual change in these variables using linear (probability) models. Differences in annual change across regions were examined by adding interaction terms and running a Wald-type test for the joint significance of interaction terms. Finally, we explored the amount of variance in outcomes that was explainable at the region level by estimating intraclass correlation coefficients (ICCs) from random-intercept linear models that nested participants in regions for SRMDs and stigma-related indicators (ICCs are detailed in Supplementary Table 3).

Analyses were done with the survey weights provided in the HSE and AMI to derive nationally representative estimates. Analyses were done in the complete-case samples of 78,226 participants (98.9% of whole sample) in the HSE and 17,287 participants (99.3% of whole sample) in the AMI with data on all variables in Stata 17 and R 4.1.2. Models in the HSE dataset were reproduced to show annual relative change, using prevalence ratios from Poisson regression, in Supplementary Table 4.

## 3. RESULTS

### 3.1 Self-reported mental disorders (SRMDs) across English regions

Table 1 presents the prevalence of SRMDs in 2009 and 2018, and annual changes in prevalence between 2009-18, at the national level and across the nine regions. Figure 1 presents time trends in the prevalence of SRMDs across regions between 2009-18.

**TABLE 1.** Time trends in self-reported mental disorders in England, ages 16+. Health Survey for England (HSE), 2009-18 (n = 78,226)

|  |  | 2009<br>N = 4,618 |  | 2018<br>N = 8,052 |  | Annual absolute change<br>(in percentage points) |  |  |  |
| --- | --- | --- | --- | --- | --- | --- | --- | --- | --- |
|  |  | % | 95%CI | % | 95%CI | Bivariate |  | Adjusted* |  |
|  |  |  |  |  |  | B | 95%CI | B | 95%CI |
|  | Average | 4.3 | 3.6-5.0 | 9.1 | 8.3-9.9 | **0.6** | **0.5-0.7** | **0.6** | **0.5-0.7** |
| 1 | North East | 6.1 | 3.5-10.3 | 11.9 | 9.3-15.0 | **0.9** | **0.6-1.1** | **0.9** | **0.6-1.2** |
| 2 | North West | 4.4 | 2.9-6.7 | 9.3 | 7.1-12.2 | **0.6** | **0.4-0.8** | **0.6** | **0.4-0.8** |
| 3 | Yorkshire & the Humberlands | 4.2 | 2.9-6.1 | 8.8 | 6.8-11.3 | **0.7** | **0.5-0.9** | **0.7** | **0.5-0.9** |
| 4 | East Midlands | 3.5 | 2.0-6.1 | 11.8 | 9.4-14.7 | **0.7** | **0.4-1.0** | **0.7** | **0.5-1.0** |
| 5 | West Midlands | 3.6 | 2.0-6.2 | 11.1 | 8.1-15.0 | **0.7** | **0.4-1.0** | **0.7** | **0.5-1.0** |
| 6 | East of England | 4.2 | 2.7-6.4 | 8.1 | 6.3-10.3 | **0.5** | **0.3-0.7** | **0.5** | **0.3-0.8** |
| 7 | London | 5.0 | 3.2-7.7 | 6.2 | 5.0-7.7 | **0.3** | **0.2-0.5** | **0.4** | **0.2-0.5** |
| 8 | South East | 4.4 | 2.7-7.2 | 8.2 | 6.3-10.6 | **0.5** | **0.3-0.8** | **0.6** | **0.3-0.8** |
| 9 | South West | 3.5 | 2.2-5.6 | 10.2 | 7.7-13.5 | **0.7** | **0.5-1.0** | **0.8** | **0.5-1.0** |
|  | Difference (p) | .916 |  | **.001** |  | **.024** |  |  |  |
|  | Difference, adjusted* (p) | .874 |  | **.013** |  |  |  | **.025** |  |

P-values represent Wald-type tests for the joint significance of 1) region dummy terms and 2) region by year interaction dummy terms based on linear probability models. Adjusted models also include age, sex, ethnicity, marital status, and social class of the person responsible for the household. Bolded estimates are statistically significant at the .05 level.

**FIGURE 1.**
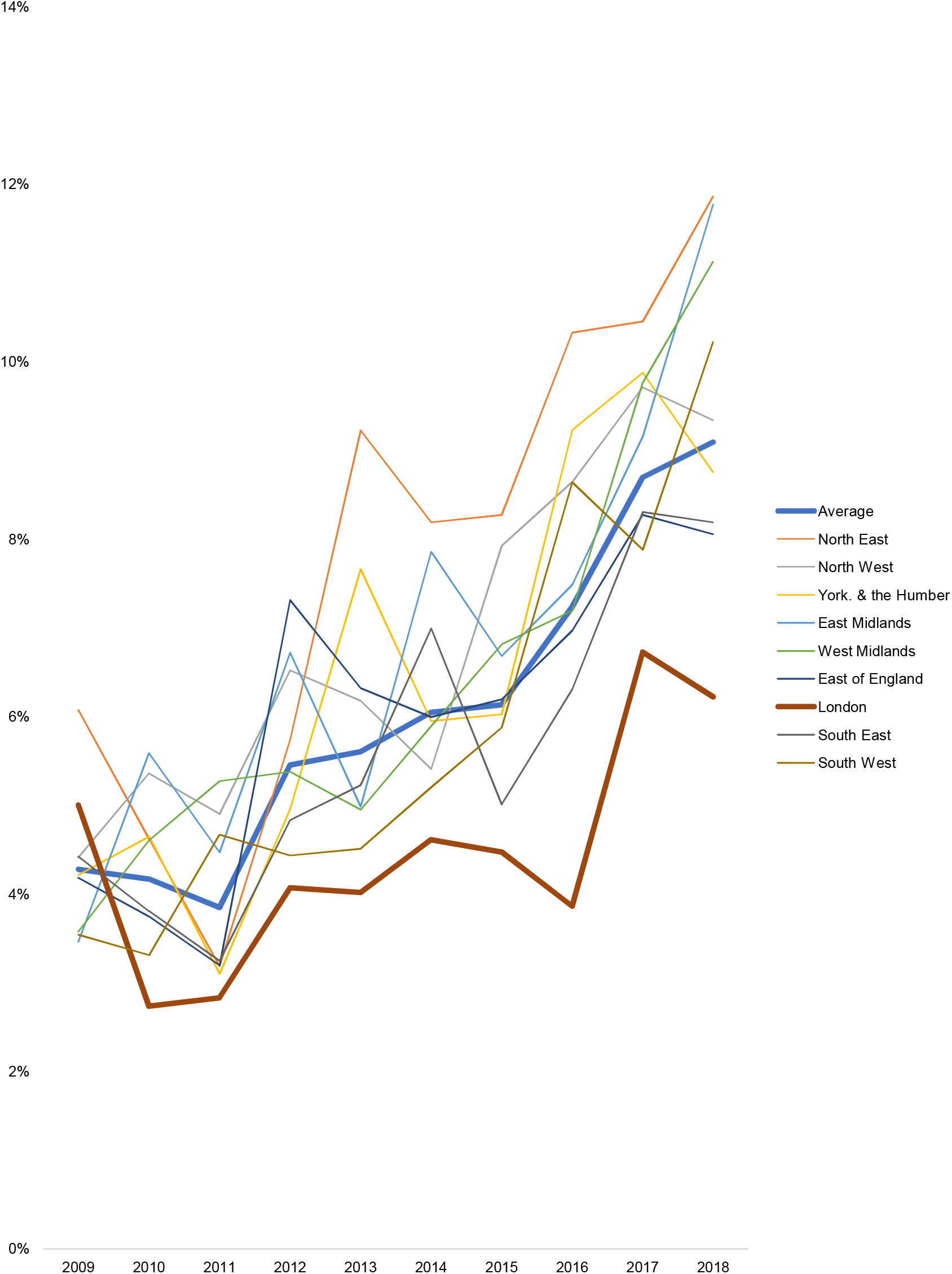
Time trends in self-reported mental disorders (%) across English regions, ages 16+. Health Survey for England, 2009-18 (n = 78,226).

The overall prevalence of SRMDs in those aged 16+ in England increased from 4.3% in 2009 to 9.1% in 2018. The prevalence of SRMDs did not vary much by region in 2009, but did in 2018. In 2009, the prevalence of SRMDs varied from a low of 3.5% in the East Midlands and South West regions to a high of 6.1% in the North East region (unadjusted *p* = .916, adjusted *p* = .874). In 2018, the prevalence of SRMDs varied between 6.2% in the London region to 11.9% in the North East region (unadjusted *p* = .001, adjusted *p* = .013).

Comparing trends across regions, whereas the annual change rate was significant in all regions, there was significant variability across them (*p* = .024): the annual absolute increase in percentage points was lowest in the London region (0.3 p.p.) and highest in the North East (0.9 p.p.). Adjusting for the five demographic covariates did not meaningfully affect these results.

### 3.2 Stigma-related indicators across English regions

Tables 2 to 4 present the average scores for stigma-related indicators in 2009 and 2017-19, and annual change rates between 2009-19, at the national level and across the nine regions. Supplementary Figure 2 presents the time trends in stigma-related indicators across regions between 2009-19.

**TABLE 2.** Time trends in <u>mental health knowledge</u> (MAKS) in England, ages 16+. Attitudes towards Mental Illness (AMI) survey, 2009-19 (n = 17,287)

|  |  | 2009<br>N = 1,743 |  | 2017-19<br>N = 3,417 |  | Annual absolute change |  |  |  |
| --- | --- | --- | --- | --- | --- | --- | --- | --- | --- |
|  |  |  |  |  |  | Bivariate |  | Adjusted* |  |
|  |  | Mean | 95%CI | Mean | 95%CI | B | 95%CI | B | 95%CI |
|  | Average | 22.2 | 22.1, 22.4 | 23.0 | 22.9, 23.1 | 0.11 | 0.09, 0.12 | 0.11 | 0.09, 0.12 |
| 1 | North East | 22.9 | 22.2, 23.6 | 23.0 | 22.3, 23.5 | 0.05 | -0.03, 0.12 | 0.06 | -0.02, 0.13 |
| 2 | North West | 22.1 | 21.7, 22.5 | 23.1 | 22.8, 23.4 | 0.15 | 0.10, 0.19 | 0.15 | 0.11, 0.19 |
| 3 | Yorkshire & the Humberlands | 22.4 | 21.9, 23.0 | 23.5 | 23.1, 23.9 | 0.12 | 0.07, 0.18 | 0.11 | 0.06, 0.17 |
| 4 | East Midlands | 22.3 | 21.8, 22.9 | 23.1 | 22.7, 23.4 | 0.08 | 0.02, 0.13 | 0.08 | 0.02, 0.13 |
| 5 | West Midlands | 22.3 | 21.8, 22.8 | 22.7 | 22.4, 23.0 | 0.07 | 0.02, 0.12 | 0.06 | 0.01, 0.10 |
| 6 | East of England | 22.8 | 22.3, 23.2 | 23.3 | 23.0, 23.6 | 0.10 | 0.05, 0.15 | 0.10 | 0.06, 0.15 |
| 7 | London | 21.7 | 21.4, 22.0 | 22.3 | 22.0, 22.6 | 0.10 | 0.06, 0.14 | 0.07 | 0.03, 0.11 |
| 8 | South East | 22.0 | 21.6, 22.3 | 23.2 | 22.9, 23.4 | 0.14 | 0.10, 0.18 | 0.14 | 0.11, 0.18 |
| 9 | South West | 22.5 | 22.0, 23.0 | 23.3 | 22.9, 23.7 | 0.13 | 0.08, 0.19 | 0.13 | 0.08, 0.19 |
|  | Difference (p) | .007 |  | < .001 |  | .106 |  |  |  |
|  | Difference, adjusted* (p) | .025 |  | < .001 |  |  |  | .007 |  |

The 6-item MAKS score can vary from 6 to 30; the observed range was 10 to 30. A higher score indicates more knowledge.
As a reference point, the weighted mean was 22.6 and weighted standard deviation was 3.2 in the full complete-case sample.
P-values represent Wald-type tests for the joint significance of 1) region dummy terms and 2) region by year interaction dummy terms based on linear probability models. Adjusted models include age, sex, ethnicity, marital status, and social class of the person responsible for the household.

**TABLE 3.** Time trends in <u>attitudes towards mental illness</u> (CAMI) in England, ages 16+. Attitudes towards Mental Illness (AMI) survey, 2009-19 (n = 17,287)

|  |  | 2009<br>N = 1,743 |  | 2017-19<br>N = 3,417 |  | Annual absolute change |  |  |  |
| --- | --- | --- | --- | --- | --- | --- | --- | --- | --- |
|  |  | Unadjusted |  | Unadjusted |  | Bivariate |  | Adjusted* |  |
|  |  | Mean | 95%CI | Mean | 95%CI | B | 95%CI | B | 95%CI |
|  | **Average** | 105.5 | 104.8, 106.2 | 110.2 | 109.7, 110.6 | **0.56** | **0.49, 0.64** | **0.57** | **0.50, 0.63** |
| **1** | **North East** | 113.5 | 110.5, 116.4 | 110.7 | 108.6, 112.7 | -0.16 | -0.47, 0.15 | -0.03 | -0.33, 0.27 |
| **2** | **North West** | 105.4 | 103.6, 107.2 | 112.1 | 110.8, 113.4 | **0.79** | **0.60, 0.98** | **0.84** | **0.66, 1.02** |
| **3** | **Yorkshire & the Humberlands** | 106.5 | 104.3, 108.8 | 111.1 | 109.6, 112.7 | **0.29** | **0.06, 0.52** | **0.25** | **0.03, 0.47** |
| **4** | **East Midlands** | 104.7 | 102.5, 107.0 | 111.2 | 109.5, 112.9 | **0.65** | **0.40, 0.91** | **0.59** | **0.35, 0.83** |
| **5** | **West Midlands** | 105.2 | 103.2, 107.2 | 110.3 | 108.9, 111.6 | **0.60** | **0.39, 0.80** | **0.57** | **0.38, 0.76** |
| **6** | **East of England** | 109.5 | 107.5, 111.5 | 110.1 | 108.6, 111.6 | 0.14 | -0.07, 0.35 | **0.23** | **0.03, 0.43** |
| **7** | **London** | 97.7 | 95.8, 99.5 | 104.5 | 103.3, 105.8 | **0.88** | **0.69, 1.06** | **0.70** | **0.52, 0.87** |
| **8** | **South East** | 106.1 | 104.5, 107.8 | 111.7 | 110.4, 112.9 | **0.75** | **0.58, 0.93** | **0.81** | **0.65, 0.98** |
| **9** | **South West** | 107.8 | 105.6, 110.0 | 112.1 | 110.6, 113.5 | **0.56** | **0.34, 0.78** | **0.58** | **0.37, 0.80** |
|  | **Difference (p)** | **< .001** |  | **< .001** |  | **< .001** |  |  |  |
|  | **Difference, adjusted\* (p)** | **< .001** |  | **< .001** |  |  |  | **< .001** |  |

The 27-item CAMI score can vary from 27 to 135; the observed range was 35 to 135. A higher score indicates less stigmatizing attitudes. As a reference point, the weighted mean was 107.8 and weighted standard deviation was 14.0 in the full complete-case sample.
P-values represent Wald-type tests for the joint significance of 1) region dummy terms and 2) region by year interaction dummy terms based on linear probability models. Adjusted models include age, sex, ethnicity, marital status, and social class of the person responsible for the household.

**TABLE 4.**
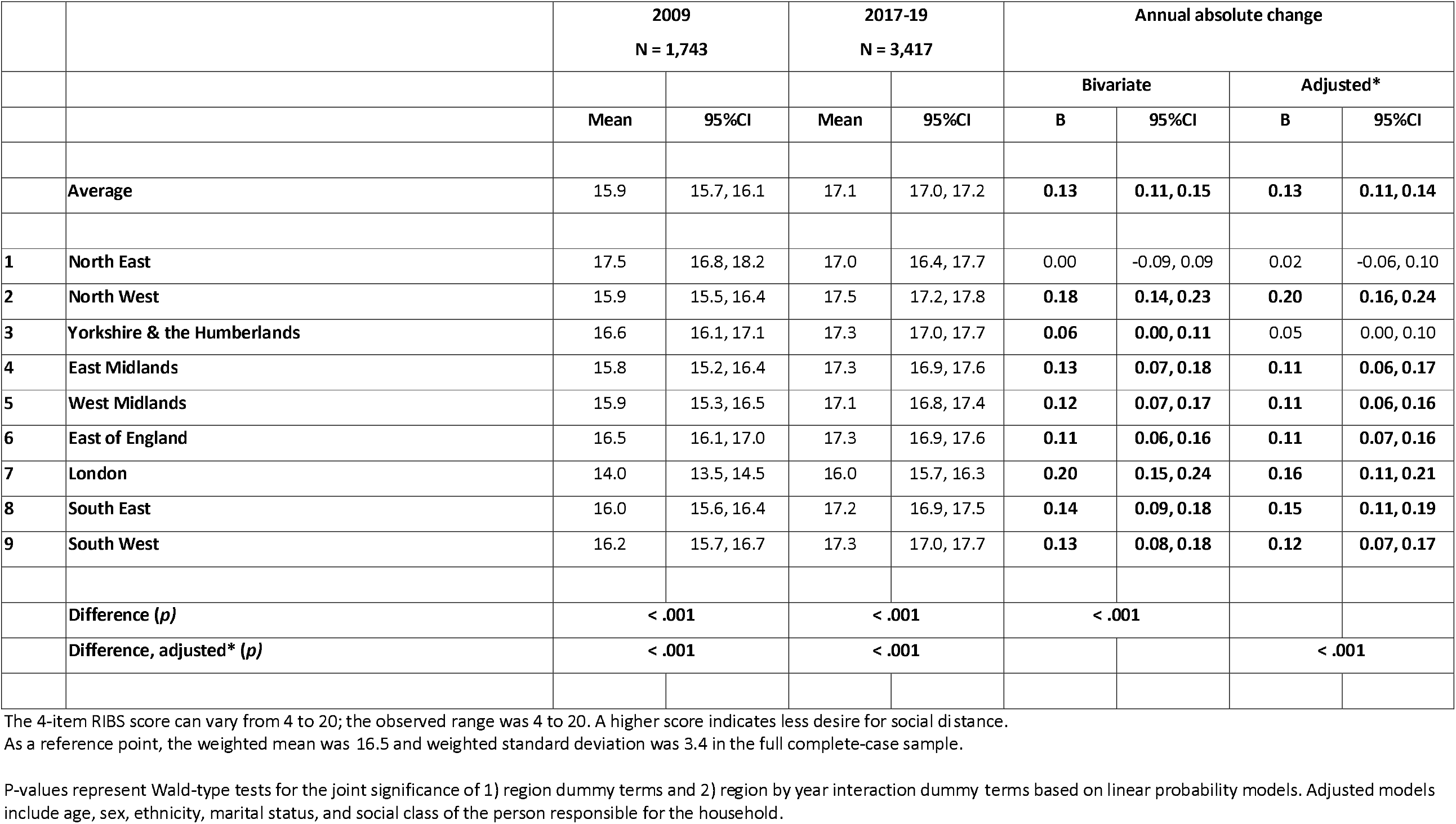
Time trends in <u>desire for social distance</u> (RIBS) in England, ages 16+. Attitudes towards Mental Illness (AMI) survey, 2009-19 (n = 17,287)

All three indicators significantly varied across regions in 2009 and 2017-19. In 2009, mean scores were each lowest in London (MAKS = 21.7, CAMI = 97.7, RIBS = 14.0) and highest in the North East region (MAKS = 22.9, CAMI = 113.5, RIBS = 17.5). Using standard deviations (SD) from the 2009 sample as a reference point, differences between these two regions represented a 0.39 SD difference for the MAKS, 1.10 SD for the CAMI, and 0.97 SD for the RIBS.

In 2017-19, scores were still lowest in London (MAKS = 22.3, CAMI = 104.5, RIBS = 16.0). The North East region, however, no longer had the most positive indicator scores; Yorkshire and the Humber reported the highest knowledge score (MAKS = 23.5) and North West reported the highest attitude score (CAMI = 112.1) and intended behaviour score (RIBS = 17.5). Using standard deviations in the 2017-19 sample as a reference point, differences between London and regions with the most positive scores represented a 0.39 SD difference for the MAKS, 0.56 SD for the CAMI, and 0.44 SD for the RIBS.

Examining change over time, all three indicators – i.e., MAKS (B = 0.11, 95%CI 0.09, 0.12), CAMI (B = 0.56, 95%CI 0.49, 0.64), and RIBS (B = 0.13, 95%CI 0.11, 0.15) – showed a significant annual improvement rate at the national level. Using standard deviations from the full complete-case sample as a reference point, average differences between 2007 and 2019 could be likened to a 0.41 SD increase for the MAKS, 0.48 SD for the CAMI, and 0.46 SD for the RIBS.

As with SRMDs, annual change rates significantly varied across regions for all indicators in adjusted models (*p* < .01). **MAKS**. In unadjusted models, MAKS scores improved most rapidly in the North West (B = 0.15, 95%CI 0.10, 0.19) and did not improve in one region: North East (B = 0.05, 95%CI - 0.03, 0.12). There were no change in statistical significance for regions’ change rate between unadjusted and adjusted models. London had a relatively small improvement rate for knowledge (adjusted model: B = 0.07, 95%CI 0.03, 0.11). **CAMI**. In unadjusted models, CAMI scores improved most rapidly in London (B = 0.56, 95%CI 0.49, 0.64) and did not improve in two regions: North East (B = -0.16, 95%CI -0.47, 0.15) and East of England (B = 0.14, 95%CI -0.07, 0.35). In adjusted models, the annual change rate in East of England increased and became significant (B = 0.23, 95%CI 0.03, 0.43). **RIBS**. In unadjusted models, RIBS scores improved most rapidly in London (B = 0.20, 95%CI 0.15, 0.24) and did not improve in one region: North East (B = 0.00, 95%CI -0.09, 0.09). In adjusted models, a second region – Yorkshire & the Humber – no longer showed a significant improvement (B = 0.05, 95% 0.00-0.10).

### 3.3 Examining variation in regional differences with intraclass correlation coefficients

Finally, we explored the extent to which regional differences in SRMDs and stigma-related indicators may be explained by contextual or compositional factors using multilevel modelling. Using unadjusted models only including dummy terms for year in the pooled HSE and AMI samples, intraclass correlation coefficients for the region level were very low for SRMDs (0.1%) and MAKS (0.8%), CAMI (3.6%), and RIBS (2.1%) scores. This supports the idea that contextual effects (including stigma at the regional level) are unlikely to be driving differences in SRMDs across regions.

## 4. DISCUSSION

This paper explored the extent to which self-reporting practices may have been sensitive to changes in public stigma by comparing time trends in self-reported mental disorders and attitudes across English regions. We provide new evidence corroborating the worrying pace at which mental health problems have increased in prevalence across English regions. Our statistical adjustment strategy supports the idea that time trends have been insensitive to changes in ethnicity and some living arrangements (i.e., marital status and social class at the household level) during this period. Multilevel modelling also supports that these differences are likely to result from regions’ composition than from contextual effects specific to the regional scale. Importantly, in keeping with austerity responses made by the English government after the 2008 Great Recession, these regional differences may result from cuts in health care and other social services across local authorities within regions [22].

### 4.1 Lack of support for the “self-report bias” hypothesis

Public stigma towards mental illness has been unequally distributed across English regions for a long period of time. Supporting the body of work using the AMI survey, we found that improvements in stigma-related indicators between 2009 and 2019 substantially varied across regions. North West, London, and the South East showed the most improvements whereas North East and the Yorkshire & Humber showed the least improvements (or no change at all). Notably, the annual change rate across indicators in London was attenuated by 20-30% when adjusting for covariates, supporting the hypothesis that improvements in this region could be explained in part by changes in socio-demographic composition over time.

These trends largely failed to support the “self-report bias” hypothesis, which predicted a corresponding increase in the prevalence of self-reported mental disorders. Instead, London showed the lowest increase in SRMDs across the nine regions and the North West and South East showed an increase matching the national average. At the other end, the North East showed the largest increase in SRMDs across regions and the Yorkshire & Humber showed a slightly above-average increase. We explore three reasons that may explain this negative finding.

The first concerns the nature of the relationship between public stigma and self-reporting, and the idea that substantial levels of improvement may be required before it can start having an impact. The role of attitudes in behaviour change is likely to be non-linear for many behaviours, and this may also be the case for the perception of shared attitudes in one’s environment [23]. While London showed above-average improvements over time, it remained in 2019 behind where most regions were in terms of attitudes and intended behaviours in 2009. It is possible that it is only once a region has a sufficiently high proportion of inhabitants with positive attitudes that it starts seeing a shift in self-reporting practices. Regions such as London may have yet to reach this point. Psychological literature also suggests that attitudes have a larger impact once they become stable over time [24]. Therefore, it is also possible that changes in the perception of shared attitudes have a lagged effect, and that the changes observed in some English regions will have an impact on self-reporting in the years to come.

A second issue concerns the scale of analysis. The use of the nine regions was done out of convenience given the design and sample sizes of the HSE and AMI surveys. However, the stigma-related processes that influence practices such as self-reporting may work at more interpersonal levels. For instance, one study found in the HSE that knowing someone with a mental health problem was associated with better help seeking practices [25]. Supporting this, another study also found in the HSE no variance in attitudes at the local area level (across geographical units averaging 1,500 inhabitants), but a meaningful degree of variance at the household level [26]. Changes in interpersonal stigma (e.g., parents and other family members having more positive attitudes over time) may therefore have a more meaningful impact on self-reporting compared with the changes in public stigma examined here.

A third reason may be that public stigma has a larger impact on self-reporting in some groups, and that our analyses failed to assess this heterogeneity. A small US study among older adults found that perceived public stigma had a stronger impact in those living in rural areas compared with urban areas [27]. In another analysis of the AMI dataset, researchers found that familiarity of someone with a mental health problem had a different impact across socioeconomic groups: compared with those in a higher social class, those in a routine job or unemployed were more likely to have positive attitudes and less desire for social distance if they knew someone with a problem or had a problem themselves [28]. Whereas the sample size of the AMI precludes us from reliably stratifying within regions (e.g., test the three-level interaction between time, region, and urbanicity), it is possible that improvements in public stigma in regions such as London have had a meaningful impact on self-reporting in some groups.

### 4.2 Towards disentangling the relationship between stigma and self-reporting

To the best of our knowledge, this study represents one of the few attempts to examine the role of changes in stigma on the self-reporting of mental health problems over time. This research programme, however, requires a range of studies to fully test this.

A first step includes using other analytic scales to examine how interpersonal and structural stigma may operate to influence self-reporting. Since our findings did not support using region as a meaningful scale, relevant scales may be more micro (i.e., households or networks) or macro (i.e., countries) [29]. Surveys using a household design with information on attitudes and mental health, such as the 2014 HSE, may offer additional insight in the role of peers’ attitudes, including parents and partners, in the self-reporting of mental health problems. On the other end, the role of structural stigma can be examined by looking at cross-country differences. Most studies have relied on the analysis of traditional media to assess national trends in public stigma, which could be contrasted with trends in self-reported mental health outcomes [30, 31]. The same could be done with social media, which is also likely to differ across countries [32, 33]. The estimation of changes in attitudes based on surveys is largely unavailable in other countries, yet cross-national comparisons support the potential for variability [34]. To support this work, other countries need to develop appropriate survey programs to assess changes in attitudes over time. Examples include data collection done between 2007 and 2013 in the U.S. Behavioral Risk Factor Surveillance System, and in 1996, 2006, and 2018 in the U.S. General Social Survey [10, 11].

### 4.3 Strength and limitations

We remind that with an ecological design (i.e., comparing aggregate levels of self-reported problems and stigma-related indicators across regions), we cannot confirm that associations at the region level match those at the individual level [35]. A cross-sectional design also precludes us from drawing causal interpretations. Reflecting on temporal ordering, it may be increases in psychiatric morbidity that are driving improvements in public stigma over time. Since mental health is a multi-faceted concept, our findings are limited by the use of only one mental health outcome, which was the only variable measured consistently across HSE waves between 2009 and 2018. Finally, the decision to use similar covariates across the HSE and AMI surveys may have led us to omit important covariates for correctly modelling change over time.

### 4.2 Conclusion

Understanding the decline in population levels of mental health, its drivers, and the policies that may tackle them represent a clear public health priority. The magnitude of this decline has led many to consider changes in public stigma over time – “de-stigmatisation” – as an alternative explanation. We shed some light on this issue by comparing time trends in self-reported mental disorders and stigma-related indicators across English regions, which provided no support for this hypothesis. Whereas public stigma and self-reporting behaviour are likely to be related, this should not be used to downplay this public health crisis.

## Data Availability

The HSE dataset used here is available on the UK Data Service platform. The AMI dataset used here is available upon reasonable request to the authors.

https://ukdataservice.ac.uk/

## Author contribution statement

TG designed the study, analysed the data, interpreted the findings, and wrote the first draft. CH provided access to the AMI dataset. CH and AM contributed to the analytic strategy, the interpretation of findings, and the writing of the paper. All authors contributed to the final version of the manuscript.

## Data availability statement

The HSE dataset was accessed on the UK Data Service platform.

