## Supplementary Material for "Is the self-reporting of mental health problems sensitive to public stigma towards mental illness? A comparison of time trends across English regions (2009-19)"

**Title**

**Last updated**

June 27^th^, 2022

**Table of contents**

- Figure 1: Stigma-related indicators in the AMI survey.
- Figure 2: Map of English regions
- Table 1: Conditions included in the HSE coding frame for self-reported mental disorders
- Table 2: Item labels for indicators in the AMI survey
- Table 3: HSE estimates, with annual relative changes.
- Table 4: ICCs for region-level variation in outcomes.

**SUPPLEMENTARY FIGURE 1**

**Stigma-related indicators across English regions, ages 16+. AMI survey, 2009-19.**

**Panel A: Knowledge (MAKS)**

|  | Average | NE | NW | Y&H | Emid | Wmid | Eeng | LDN | SE | SW |
| --- | --- | --- | --- | --- | --- | --- | --- | --- | --- | --- |
| 2009 | 22.2437 | 22.8957 | 22.1176 | 22.4405 | 22.3295 | 22.2754 | 22.7553 | 21.6949 | 21.9636 | 22.517 |
| 2010 | 22.115 | 22.7395 | 21.608 | 22.8231 | 22.6123 | 22.0857 | 22.1252 | 21.6936 | 22.2063 | 21.8609 |
| 2011 | 22.1841 | 22.0041 | 22.0113 | 22.3798 | 22.5578 | 22.2822 | 22.6181 | 21.4903 | 22.2039 | 22.3441 |
| 2012 | 22.3741 | 23.2302 | 22.4659 | 22.3525 | 22.6708 | 22.6519 | 22.3825 | 21.2919 | 22.4159 | 22.7138 |
| 2013 | 22.407 | 22.1707 | 22.643 | 22.3888 | 22.0548 | 22.4962 | 21.9652 | 21.8979 | 22.7993 | 23.0018 |
| 2014 | 22.7295 | 22.873 | 22.9618 | 22.748 | 22.779 | 22.9208 | 22.7625 | 22.1881 | 22.706 | 22.8673 |
| 2015 | 22.8789 | 23.0825 | 22.9429 | 23.3622 | 23.1371 | 23.0201 | 22.3343 | 21.9673 | 23.116 | 23.5872 |
| 2016 | 22.8569 | 23.155 | 22.8762 | 23.3426 | 22.7407 | 22.4924 | 22.7606 | 22.3135 | 22.93 | 23.3716 |
| 2017 | 22.9346 | 23.1185 | 23.2876 | 23.8501 | 22.8029 | 22.4172 | 22.8137 | 22.2574 | 22.9761 | 23.4396 |
| --- | 23.0232 | 23.0151 | 23.1432 | 23.5562 | 23.0362 | 22.7247 | 23.2729 | 22.3308 | 23.1811 | 23.2728 |
| 2019 | 23.1117 | 22.9117 | 22.9988 | 23.2623 | 23.2695 | 23.0322 | 23.7322 | 22.4042 | 23.3862 | 23.106 |

**Panel B: Attitudes (CAMI)**

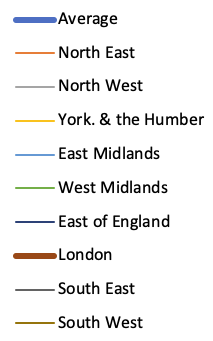

|  | Average | NE | NW | Y&H | Emid | Wmid | Eeng | LDN | SE | SW |
| --- | --- | --- | --- | --- | --- | --- | --- | --- | --- | --- |
| 2009 | 105.506 | 113.453 | 105.378 | 106.51 | 104.707 | 105.23 | 109.493 | 97.658 | 106.127 | 107.782 |
| 2010 | 106.145 | 113.379 | 105.765 | 109.163 | 107.844 | 105.212 | 108.53 | 99.5375 | 106.229 | 106.683 |
| 2011 | 105.746 | 105.865 | 105.254 | 111.115 | 106.031 | 105.872 | 108.712 | 98.7193 | 104.112 | 109.529 |
| 2012 | 106.375 | 111.921 | 106.615 | 110.229 | 105.808 | 108.82 | 106.858 | 97.8225 | 105.845 | 109.369 |
| 2013 | 107.129 | 105.776 | 108.377 | 108.036 | 106.375 | 107.232 | 106.813 | 101.716 | 109.65 | 109.794 |
| 2014 | 108.594 | 108.487 | 110.872 | 111.395 | 108.657 | 109.75 | 108.96 | 101.476 | 108.787 | 110.777 |
| 2015 | 108.93 | 111.103 | 111.497 | 111.044 | 109.21 | 107.12 | 105.262 | 103.945 | 110.12 | 111.806 |
| 2016 | 109.685 | 110.79 | 109.203 | 111.001 | 110.526 | 108.577 | 109.304 | 106.891 | 109.528 | 113.359 |
| 2017 | 109.939 | 113.272 | 113.43 | 113.502 | 111.338 | 107.267 | 108.675 | 103.712 | 110.414 | 113.104 |
| --- | 110.153 | 110.707 | 112.071 | 111.458 | 111.19 | 110.453 | 110.035 | 104.553 | 111.707 | 111.986 |
| 2019 | 110.367 | 108.142 | 110.711 | 109.414 | 111.043 | 113.638 | 111.394 | 105.394 | 112.999 | 110.869 |

**Panel C: Behaviour (RIBS)**

|  | Average | NE | NW | Y&H | Emid | Wmid | Eeng | LDN | SE | SW |
| --- | --- | --- | --- | --- | --- | --- | --- | --- | --- | --- |
| 2009 | 15.8851 | 17.4702 | 15.9255 | 16.6269 | 15.8224 | 15.903 | 16.5315 | 14.0321 | 15.9907 | 16.1654 |
| 2010 | 16.1239 | 16.589 | 16.1242 | 16.9558 | 16.6973 | 16.0043 | 16.2346 | 15.0234 | 16.1894 | 16.1072 |
| 2011 | 15.962 | 15.8167 | 16.0699 | 16.751 | 15.9096 | 16.3084 | 16.1902 | 14.4056 | 16.0558 | 16.4665 |
| 2012 | 16.1625 | 16.9592 | 15.7592 | 17.0549 | 16.4865 | 16.9366 | 16.277 | 14.7993 | 15.9869 | 16.3274 |
| 2013 | 16.3945 | 16.6188 | 17.2432 | 16.6172 | 16.4208 | 16.0459 | 16.1586 | 15.4175 | 16.7197 | 16.4251 |
| 2014 | 16.6577 | 16.7648 | 17.2522 | 17.4121 | 16.8992 | 17.0633 | 16.4908 | 15.4256 | 16.5036 | 16.5916 |
| 2015 | 16.6335 | 17.134 | 16.8539 | 17.0862 | 16.6728 | 16.4225 | 16.0073 | 15.9139 | 16.8657 | 17.0849 |
| 2016 | 16.7423 | 16.6825 | 17.3874 | 17.098 | 16.872 | 16.5064 | 16.9006 | 15.9069 | 16.4345 | 17.1413 |
| 2017 | 17.1452 | 18.1091 | 17.7726 | 17.8671 | 17.3791 | 16.637 | 16.9686 | 16.074 | 16.9864 | 17.8039 |
| --- | 17.0701 | 17.0509 | 17.5142 | 17.4074 | 17.264 | 17.1197 | 17.2453 | 15.9982 | 17.2267 | 17.292 |
| 2019 | 16.995 | 15.9926 | 17.2558 | 16.9478 | 17.1489 | 17.6025 | 17.5219 | 15.9223 | 17.4669 | 16.78 |

N.B. A higher score indicates a better outcome.

**SUPPLEMENTARY FIGURE 2**

**Map of English Regions**

**East Midlands**

**West Midlands**

**South West**

**South East**

**London**

**East of England**

**North West**

**Yorkshire & the Humberlands**

**North East**

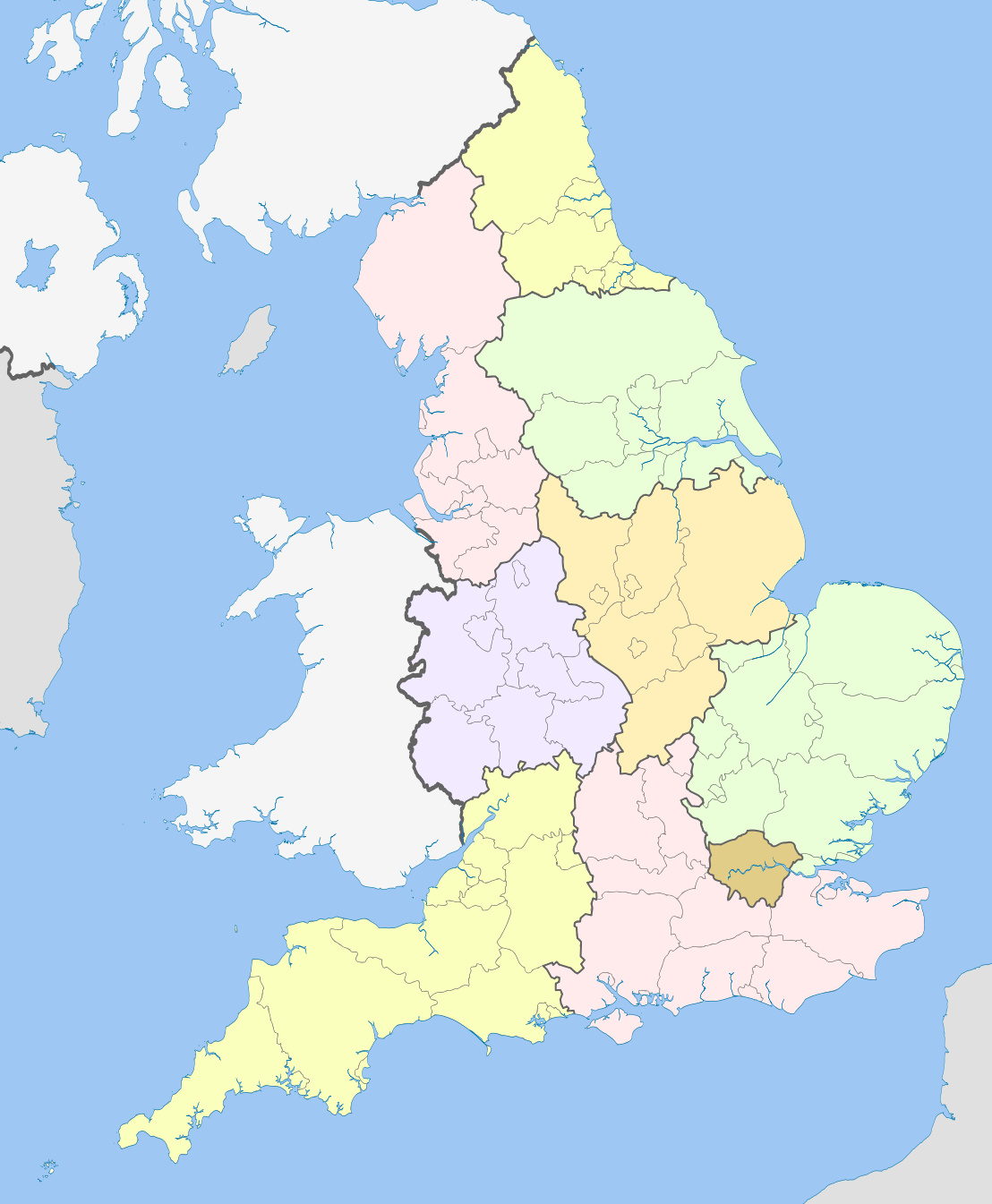

**SUPPLEMENTARY TABLE 1**

**Conditions included in the HSE coding frame for self-reported mental disorders**

**04 Mental illness/anxiety/depression/nerves (nes)**

- Alcoholism, recovered not cured alcoholic
- Angelman Syndrome
- Anorexia nervosa
- Anxiety, panic attacks
- Asperger Syndrome
- Autism/Autistic
- Bipolar Affective Disorder
- Catalepsy
- Concussion syndrome
- Depression
- Drug addict
- Dyslexia
- Hyperactive child.
- Nerves (nes)
- Nervous breakdown, neurasthenia, nervous trouble
- Phobias
- Schizophrenia, manic depressive
- Senile dementia, forgetfulness, gets confused
- Speech impediment, stammer
- Stress

**05 Mental handicap**

- Incl. Down's syndrome, Mongol
- Mentally retarded, subnormal

**SUPPLEMENTARY TABLE 2**

**Item labels for indicators in the AMI survey**

|  | **CAMI 27-item scale of community attitudes towards mental illness** |
| --- | --- |
| 1 | One of the main causes of mental illness is a lack of self- discipline and will-power |
| 2 | There is something about people with mental illness that makes it easy to tell them from normal people |
| 3 | As soon as a person shows signs of mental disturbance, he should be hospitalized |
| 4 | Mental illness is an illness like any other |
| 5 | Less emphasis should be placed on protecting the public from people with mental illness |
| 6 | Mental hospitals are an outdated means of treating people with mental illness |
| 7 | Virtually anyone can become mentally ill |
| 8 | People with mental illness have for too long been the subject of ridicule |
| 9 | We need to adopt a far more tolerant attitude toward people with mental illness in our society |
| 10 | We have a responsibility to provide the best possible care for people with mental illness |
| 11 | People with mental illness don't deserve our sympathy |
| 12 | People with mental illness are a burden on society |
| 13 | Increased spending on mental health services is a waste of money |
| 14 | There are sufficient existing services for people with mental illness |
| 15 | People with mental illness should not be given any responsibility |
| 16 | A woman would be foolish to marry a man who has suffered from mental illness, even though he seems fully recovered |
| 17 | I would not want to live next door to someone who has been mentally ill |
| 18 | Anyone with a history of mental problems should be excluded from taking public office |
| 19 | No-one has the right to exclude people with mental illness from their neighbourhood |
| 20 | eople with mental illness are far less of a danger than most people suppose |
| 21 | Most women who were once patients in a mental hospital can be trusted as babysitters |
| 22 | The best therapy for many people with mental illness is to be part of a normal community |
| 23 | As far as possible, mental health services should be provided through community based facilities |
| 24 | Residents have nothing to fear from people coming into their neighbourhood to obtain mental health services |
| 25 | It is frightening to think of people with mental problems living in residential neighbourhoods |
| 26 | Locating mental health facilities in a residential area downgrades the neighbourhood |
| 27 | People with mental health problems should have the same rights to a job as anyone else |
|  | **RIBS 4-item scale of desire for social distance from someone with a mental health problem** |
| 1 | In the future, I would be willing to live with someone with a mental health problem |
| 2 | In the future, I would be willing to work with someone with a mental health problem |
| 3 | In the future, I would be willing to live nearby to someone with a mental health problem |
| 4 | In the future, I would be willing to continue a relationship with a friend who developed a mental health problem |
|  | **MAKS 6-item scale of knowledge about mental health problems** |
| 1 | Most people with mental health problems want to have paid employment |
| 2 | If a friend had a mental health problem, I know what advice to give them to get professional help |
| 3 | Medication can be an effective treatment for people with mental health problems |
| 4 | Psychotherapy […] can be an effective treatment for people with mental health problems |
| 5 | People with severe mental health problems can fully recover |
| 6 | Most people with mental health problems go to a health care professional to get help |

**SUPPLEMENTARY TABLE 3**

**Time trends in self-reported mental disorders in England, ages 16+. Health Survey for England (HSE), 2009-18 (n = 78,226)**

|  |  | **2009**  **N = 4,618** | | **2018**  **N = 8,052** | | **Annual RELATIVE change**  **(percentage)** | | | |
| --- | --- | --- | --- | --- | --- | --- | --- | --- | --- |
|  |  |  |  |  |  | **Bivariate** | | **Adjusted*** | |
|  |  | **%** | **95%CI** | **%** | **95%CI** | **PR** | **95%CI** | **PR** | **95%CI** |
|  | **Average** | 4.3 | 3.6-5.0 | 9.1 | 8.3-9.9 | **1.10** | **1.09-1.12** | **1.11** | **1.09-1.12** |
| **1** | **North East** | 6.1 | 3.5-10.3 | 11.9 | 9.3-15.0 | **1.12** | **1.08-1.16** | **1.13** | **1.09-1.17** |
| **2** | **North West** | 4.4 | 2.9-6.7 | 9.3 | 7.1-12.2 | **1.09** | **1.05-1.13** | **1.09** | **1.06-1.13** |
| **3** | **Yorkshire & the Humberlands** | 4.2 | 2.9-6.1 | 8.8 | 6.8-11.3 | **1.11** | **1.08-1.15** | **1.12** | **1.08-1.16** |
| **4** | **East Midlands** | 3.5 | 2.0-6.1 | 11.8 | 9.4-14.7 | **1.11** | **1.07-1.16** | **1.12** | **1.07-1.16** |
| **5** | **West Midlands** | 3.6 | 2.0-6.2 | 11.1 | 8.1-15.0 | **1.12** | **1.07-1.17** | **1.12** | **1.07-1.17** |
| **6** | **East of England** | 4.2 | 2.7-6.4 | 8.1 | 6.3-10.3 | **1.09** | **1.05-1.13** | **1.09** | **1.05-1.13** |
| **7** | **London** | 5.0 | 3.2-7.7 | 6.2 | 5.0-7.7 | **1.08** | **1.04-1.12** | **1.08** | **1.04-1.13** |
| **8** | **South East** | 4.4 | 2.7-7.2 | 8.2 | 6.3-10.6 | **1.10** | **1.06-1.14** | **1.11** | **1.06-1.15** |
| **9** | **South West** | 3.5 | 2.2-5.6 | 10.2 | 7.7-13.5 | **1.14** | **1.09-1.18** | **1.15** | **1.10-1.19** |
|  | **Difference (*p)*** |  |  |  |  | .671 | |  |  |
|  | **Difference, adjusted* (*p)*** |  |  |  |  |  |  | .652 | |

Estimates represent the annual relative change in the probability of reporting a mental health problem. P-values represent Wald-type tests for the joint significance of region by year interaction dummy terms based on Poisson models. Adjusted models also include age, sex, ethnicity, marital status, and social class of the person responsible for the household. Bolded estimates are statistically significant at the .05 level.

PR = Prevalence ratio.

**SUPPLEMENTARY TABLE 4**

**Intraclass correlation coefficients (ICC) for self-reported mental disorders (SRMDs) and stigma-related indicators in the HSE and AMI surveys.**

| **Dataset** | **HSE** | | | **AMI** | | |
| --- | --- | --- | --- | --- | --- | --- |
| **Outcome** | **SRMD** | | | **CAMI** | **RIBS** | **MAKS** |
| **Weight*** | **Unscaled** | **Method A** | **Method B** |  |  |  |
| **Level** | **%** | **%** | **%** | **%** | **%** | **%** |
| **Bivariate** |  |  |  |  |  |  |
| Region | 0.1 | 0.1 | 0.1 | 3.6 | 2.1 | 0.8 |
| **Adjusted** |  |  |  |  |  |  |
| Region | < 0.1 | < 0.1 | < 0.1 | 1.8 | 1.3 | 0.6 |

ICCs can be understood as the proportion of the unexplained variance at each level. Models were random-intercept linear (probability) models in the complete-case samples. Adjusted models include age, sex, ethnicity, marital status, and social class of the person responsible for the household. Models for SRMDs in the HSE were ran without the primary sampling unit and strata variables.

* Multi-level models should not readily incorporate the survey weight made available by HSE. The three ICCs shown are produced using the unscaled and two scaled weights as proposed by Carle (2009). This was not done in the AMI as weight scaling requires information on stratification.

*Carle AC. Fitting multilevel models in complex survey data with design weights: Recommendations. BMC Med Res Methodol. 2009; 9: 49. doi: 10.1186/1471-2288-9-49.*
